# Weekly SARS-CoV-2 screening of asymptomatic students and staff to guide and evaluate strategies for safer in-person learning

**DOI:** 10.1101/2021.03.20.21253976

**Authors:** Shira Doron, Robin R. Ingalls, Anne Beauchamp, Jesse Boehm, Helen W. Boucher, Linda H. Chow, Linda Corridan, Katey Goehringer, Doug Golenbock, Liz Larsen, David Lussier, Marcia Testa, Andrea Ciaranello

**Author notes:** These authors contributed equally to this work (Shira Doron, Robin R. Ingalls). **Communicating author:** Andrea Ciaranello, MD, MPH, 100 Cambridge St., Room 1670, Boston, MA 02114.

## Abstract

**Background:** Data suggest that COVID-19 transmission in K-12 schools is uncommon, but few studies have confirmed this using widespread screening of asymptomatic individuals.

**Objective:** To evaluate the incidence of asymptomatic COVID-19, document the frequency of in-school transmission, and confirm feasibility of widespread asymptomatic screening in schools.

**Design:** Prospective observational study

**Setting:** Single mid-sized suburban school district including 10 schools and a central office.

**Participants:** District staff and students

**Interventions:** Asymptomatic screening PCR for SARS-CoV-2

**Measurements:** Concurrent with a hybrid model and layered mitigation, weekly pooled testing of staff and secondary students was offered using saliva samples collected at home. Identification of >1 case in a school prompted investigation for possible in-school transmission. Staff and families were surveyed about satisfaction with the screening program.

**Results:** From weeks 1-18, rates of incident COVID-19 in the surrounding community rose steadily. Weekly staff and student screening identified 0-7 asymptomatic cases/week. In week 7, 5 cases were identified among staff who shared an office setting. Enhancements to mitigation strategies were undertaken. The proportion of survey respondents self-reporting comfort with in-person learning before versus after implementation of screening increased.

**Limitations:** Because screening testing was not mandatory, the results from the participating population might not represent the entire school community.

**Conclusions:** In this school district with layered mitigation measures, in-school transmission was rare. The program identified a cluster with in-school staff-to-staff transmission and spurred enhancement of safety strategies. A weekly COVID-19 screening program can provide critical data to inform mitigation efforts, and provides school-specific, current data to inform decisions about in-person learning models. Screening provided reassurance and identified asymptomatic cases.

**Funding:** The Wellesley Education Foundation provided funding for the testing.

## INTRODUCTION

The spring 2020 onset of the COVID-19 pandemic led to a global transition to remote learning. Many US schools persisted in this model in 2020-21: 49% of districts^1^ and 52% of students^2^ planned remote education in September 2020, with higher rates of remote-only learning in communities of color. Recognizing the challenges and inequities of remote learning, many schools offer in-person learning with COVID-19 mitigation strategies, including masking, distancing, ventilation upgrades, reduced occupancy, and enhanced disinfection.^3,4^

Where community rates are high, it is expected that students and staff who enter school buildings will unknowingly be infected with COVID-19. A central question is whether they will transmit to others in the school setting. Many countries have kept schools open for in-person learning despite rising community COVID-19 incidence (often while restricting other sectors of society);^5,6^ data from both international and US settings suggest that where mitigation strategies are well implemented, in-school transmission of COVID-19 is rare.^7–9^ These findings are reassuring with regard to symptomatic and severe illness among students and educators/staff. However, the lack of widespread SARS-CoV-2 testing of students and faculty/staff without symptoms (“screening”) precludes these reports from assessing the true number of in-school exposures that result in asymptomatic infection, possibly placing educator/staff and student household members at risk.

Regular screening in K-12 schools may provide several benefits: information about in-school risk, a snapshot of community COVID-19 prevalence, and an additional risk mitigation strategy (by identifying and isolating people with COVID-19 before onward transmission occurs).^10^ The high cost of SARS-CoV-2 PCR testing, limited access to accurate rapid assays, and the challenges of sample collection and data management in public-school settings have limited the uptake of screening programs, especially in public-school districts with limited resources. We developed a consortium of 25 public-school districts to implement and evaluate screening programs and collectively negotiate lower screening costs. We report the results of a screening program in one well-resourced member district with the most mature screening program. With the goal of ‘advocacy through action,’ our objectives were to describe barriers and facilitators of K-12 public school screening; advocate for accurate, low-cost screening options; and facilitate rollout of similar programs in schools across the state to improve equity and access to in-person learning.^11^

## METHODS

This public-school district supports Pre-K to Grade 12 students in 1 preschool, 7 elementary schools (grades K-5), 1 middle school (grades 6-8) and 1 high school (grades 9-12). In fall 2020, families were offered the option to enroll students in a fully-remote curriculum (approximately 500 students) or a hybrid-learning curriculum (approximately 4000 students). Schools opened to remote learning for all students on September 16^th^. Hybrid learning began October 1, consisting of in-person learning 2 days per week with remote learning 2.5 days per week; students with high learning needs attended in-person 4 days per week, along with all students in Grades K, 1, and later Grade 2. Safety measures were based on the CDC and state Department of Public Health and Department of Education guidelines, and included mandatory masking (except during lunch and designated mask breaks), frequent hand sanitizing, 6 feet of distance separating students seated at their desks in most classrooms, ventilation upgrades as feasible based on age of individual buildings, use of outdoor space when possible, enhanced environmental disinfection, and daily symptom screening.^12^

Baseline individual SARS-CoV-2 PCR screening was offered to all staff and students prior to the opening of hybrid learning, followed 1 month later by weekly pooled PCR screening for all staff as well as all students in the middle and high schools. Participation in screening was not required but strongly encouraged. Parents and guardians provided consent for student testing. Saliva collection packages were assembled, labeled with unique barcodes, and distributed to staff and students at school. Students or staff who tested positive for SARS-CoV-2 within the previous 90 days were excluded from participation that week. Saliva samples (1 ml volume) were collected at home, then returned to the schools for shipping to a commercial laboratory. Screening was scheduled to interact with hybrid learning schedules; students present on Monday/Thursday were screened on Mondays, enabling result-return and contact tracing over 48 hours of remote learning, and students present Tuesday/Friday were screened on Tuesdays.

Screening for SARS-CoV-2 was performed using a pooled saliva PCR directed at two targets in the nucleocapsid gene, N1 and N2 (Saliva Clear, New York State Department of Health-authorized, US FDA emergency use authorization (EUA) pending). Samples failing to demonstrate DNA on quality testing were not assayed. If a pool of 24 tests resulted as positive, pooled testing was repeated with sequentially fewer individual samples, until positive results were isolated to a pool of 2. Those two individuals were then contacted for consent to perform individual diagnostic testing (Saliva Direct PCR, US FDA EUA), at which time US Department of Health and Human Services (HHS)-required demographic data were collected. No additional specimen was required to proceed to diagnostic testing. Students and staff from positive pairs were instructed to isolate while awaiting diagnostic test results. Positive diagnostic SARS-CoV-2 test results were reported directly to the staff and parents/guardians of students, as well as local health officials and the district director of nursing.

Confirmed SARS-CoV-2 infection was defined when an individual specimen was positive for SARS-CoV-2 by follow-up diagnostic RT-PCR. Confirmed cases were further classified as symptomatic (symptom onset before the first positive laboratory test), presymptomatic (first positive laboratory test before subsequent symptom onset), or asymptomatic (positive laboratory test but clinical criteria never met). All individuals with a positive test result were required to isolate per CDC and state Department of Public Health (DPH) guidelines.^13^

Confirmed cases resulted in contact tracing performed by the health department linked to the address of each infected individual (to identify community contacts, as resources allowed) and the school nursing department (for in-school contacts), in accordance with DPH guidelines.^13^ In-school contact tracing identified individuals with which the infected person spent at least 15 minutes within 6’ (consecutive exposure through 10/21/20, cumulative exposure thereafter), regardless of masking.^12^ The communicable period for which contact tracing was required began 2 days before symptom onset or, if no symptoms ever developed, 2 days prior to the collection date of the positive test. Close contacts were instructed to quarantine and seek testing according to state guidelines (14 days regardless of testing through 12/2/20, with options for shorter quarantine periods following negative tests thereafter).^14^

Throughout the screening program, educators/staff and students were regularly reminded to stay out of school and seek outside diagnostic testing if they developed any symptoms consistent with COVID-19; some also sought outside testing after exposures or travel.^12^ In the event that >1 case was identified in a single school over a 2-week period, either through screening or outside testing for any reason, an investigation was initiated to determine the presence or absence of in-school transmission. This investigation included review for possible exposures in and outside of school and mapping of student and educator/staff movement patterns and locations of staff workstations. Educators/staff and students also answered detailed questions regarding location and duration of mask breaks, eating, and drinking; shared objects and surfaces; and recall of any symptoms prior to detection of the cluster.

An online survey was sent to educators/staff and families/caregivers of students prior to the initiation of the screening program and repeated at screening week 11.^15^ Domains included: level of comfort with in-person schooling without baseline testing, degree of reassurance about school safety after results of baseline testing; change in comfort with in-person learning due to the screening program; and perceived benefits and detriments of weekly screening. Program staff tallied the number and type of personnel and number of hours spent by each to implement screening.

### Role of the funding source and ethical review

The Wellesley Education Foundation provided financial and programmatic support for screening, including survey design, implementation, and analysis. This study was approved by the Mass General Brigham Institutional Review Board.

## RESULTS

### Identification of COVID-19 through the screening program

A total of 921 district staff were eligible for participation in the screening program, including 256 at the high school and 258 at the middle school/central office. Most staff (87%) did not live in the town where the school district is situated. Students eligible for screening included 1,403 at the high school and 1,000 at the middle school. Excluding holiday weeks (weeks 8, 9, 14) and school closures, participation in screening varied ranged by week from 58-77% of eligible students and 73-83% of eligible staff. Common reasons for failure to submit a sample included absence from school on the day of kit distribution or collection or forgotten samples. The turnaround time from receipt of samples to results of a negative pool was approximately 48 hours; time from consent to perform diagnostic testing to verification of diagnostic results was 2-4 hours.

During the screening program period, rates of incident COVID-19 in the district’s municipality rose steadily from 5 to 32/100,000/day (Table 1). From weeks 1 to 18, 126 COVID-19 cases were identified among faculty/staff and students who were enrolled in any in-person learning model (Figure 1). Of these 126 cases, 37 were identified through the screening program, and 89 were identified through outside tests.

**Table 1.** Results of a public K-12 school screening program.

| Week | Staff (total N = 921) |  |  | Students (total N =2,403) |  |  | Town COVID-19 metrics* |  |
| --- | --- | --- | --- | --- | --- | --- | --- | --- |
|  | Participated in surveillance (n) | Positive results by surveillance (n) | Positive results by outside testing (n) | Participated in surveillance (n) | Positive results by surveillance (n) | Positive by outside testing (n) | 14-day average of daily cases/ 100K | Molecular test positivity (%) |
| 1 | 1005 | 0 | 2 | 3,596** | 1 | 0 | 2.2 | 0.09 |
| 2 |  |  | 0 |  |  | 0 | 2.4 | 0.08 |
| 3 |  |  | 0 |  |  | 0 | 2.6 | 0.10 |
| 4 | 363 | 0 | 1 |  |  | 2 | 1.7 | 0.09 |
| 5 | 721 | 0 | 0 |  |  | 0 | 3.4 | 0.17 |
| 6 | 739 | 1 | 0 | 1,847 | 0 | 0 | 5.8 | 0.26 |
| 7 | 687 | 5 | 6 | 1,648 | 2 | 1 | 5.3 | 0.23 |
| 8 | 584 | 2 | 0 | 774 | 1 | 4 | 13.7 | 0.58 |
| 9 | 602 | 2 | 0 | 759 | 0 | 7 | 19.5 | 0.78 |
| 10 | 737 | 0 | 2 | 1,774 | 2 | 2 | 19.5 | 0.88 |
| 11 | 749 | 0 | 1 | 1,636 | 3 | 2 | 19.7 | 0.98 |
| 12 | 763 | 0 | 1 | 1,578 | 2 | 8 | 20.9 | 1.05 |
| 13 | 735 | 0 | 4 | 1,387 | 0 | 8 | 21.6 | 1.26 |
| 14 | 0 | 0 | 0 | 0 | 0 | 0 | 18.8 | 1.58 |
| 15 | 727 | 1 | 2 | 1,576 | 6 | 10 | 19.5 | 2.34 |
| 16 | 673 | 1 | 3 | 1,579 | 3 | 4 | 24.5 | 2.11 |
| 17 | 722 | 1 | 1 | 1,622 | 2 | 11 | 25.0 | 1.61 |
| 18 | 733 | 1 | 1 | 733 | 3 | 4 | 31.5 | 1.59 |

\* From Massachusetts Department of Public Health.<sup>46</sup> 87% of school staff live in other towns.
\*\*Baseline screening was offered to students at all grade levels (preK-12). Subsequent screening (weeks 6-18) was offered to middle and high school students (n=2,403).

**Figure 1.**
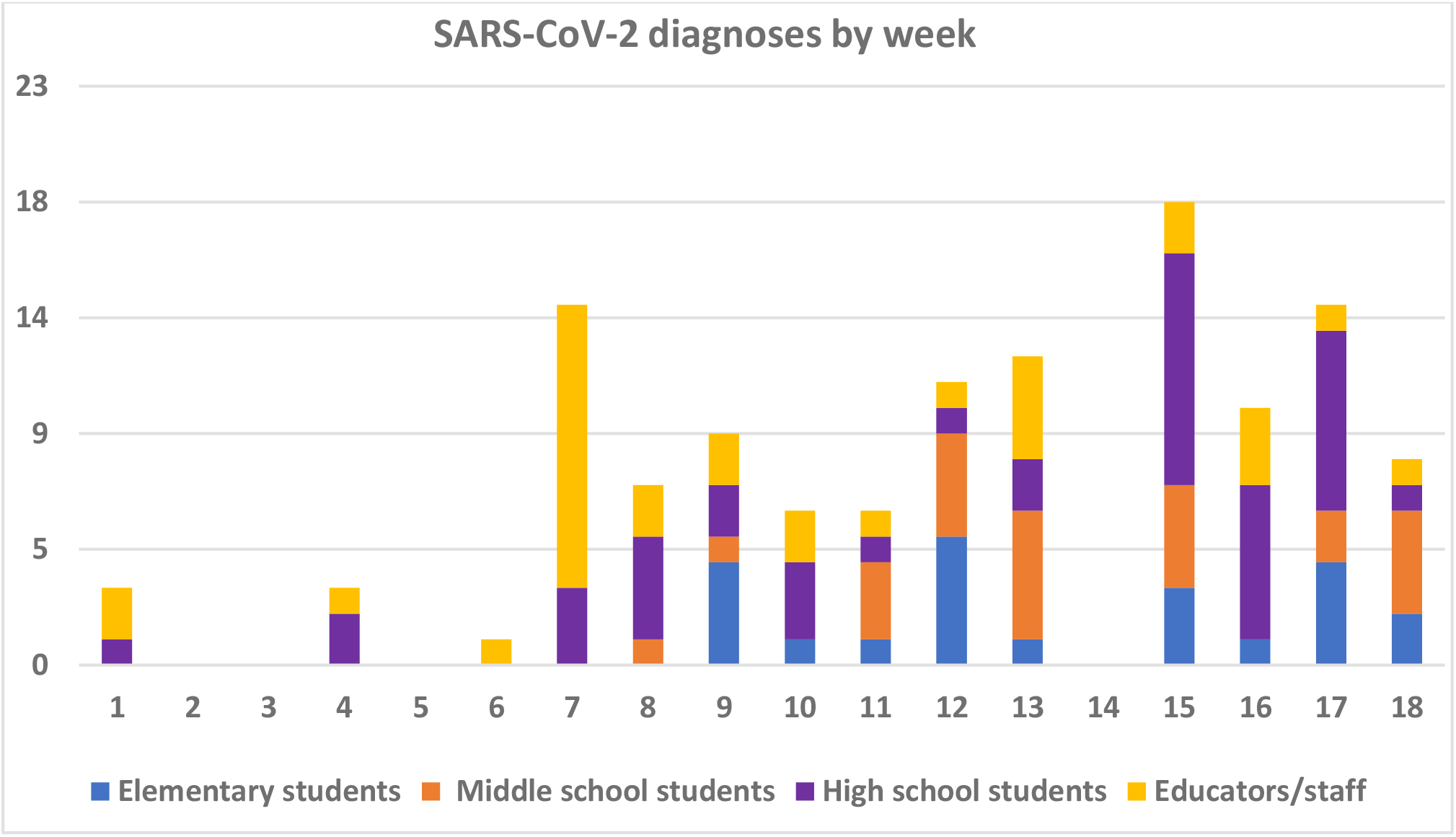
SARS-CoV-2 diagnoses during 18 weeks of in-person learning. The horizontal axis shows week number. The vertical axis shows number of confirmed SARS-CoV-2 diagnoses among members of the school community for students (stratified by grade level) and educators/staff.

In week 6, screening identified 1 positive staff member in the high school main office. Three additional main office staff members were identified as close contacts; all sought individual testing during quarantine and tested positive. In week 7, 4 additional main office staff members not identified as close contacts had positive results from the weekly screening, as well as 1 staff member not based in the main office and 2 high school students. Based on these results, the high school was transitioned to remote learning for a period of 3 weeks, to allow for full investigation. Possible risk factors for transmission and opportunities for reinforcement of existing protocols were identified in three categories. First, review of heating, ventilation, and air conditioner (HVAC) systems, including assessment of airflow in the main office using smoke, suggested airflow impedance in the main office area attributed to plexiglass dividers with sidewalls; these structures were rearranged. Second, location mapping and staff interviews highlighted occasions of mask removal at distances <6’, primarily for eating and drinking; additional spaces for unmasking and eating/drinking were provided. Third, high-traffic areas (especially in two-way entry/egress areas) and shared offices were identified; one-way traffic flow and schedules for single office occupancy were established. Because no additional cases were identified in screening among staff at other schools, or among middle school students, all other schools in the district continued hybrid or in-person learning.

### Identification of COVID-19 outside the screening program

In week 9, 3 cases were identified in one elementary school classroom, all through outside testing of close contacts. After evaluation by school and board of health officials, transmission was thought to have occurred outside of school. This single class moved to remote learning for 10 calendar days (including a professional development day, thus missing one full school week). Screening at the middle and high school and among elementary staff revealed few additional cases, and in-person learning continued at other sites. In week 15 (post-holiday), 9 students with SARS-CoV-2 were identified at the high school: 5 via the screening program and 4 via outside testing. The high school transitioned to remote learning for 10 calendar days (including 2 weekends, thus missing one full school week), and in-person learning remained in place at other schools.

### Implementation and acceptability

Survey responses were received from 491 educators/staff and 658 families/caregivers (78% with at least one student at middle or high school levels, 22% with students only in preK-5), a response rate of 53% (educators/staff) and 12-24% (families/caregivers, assuming 1-2 responses per family). Prior to baseline testing, the proportion of educators/staff who reported feeling “mostly comfortable” or “very comfortable” was 12%; after baseline testing, 82% reported feeling “reassured” or “very reassured” (Table 2). Among families/caregivers, these proportions were 39% and 87%.

**Table 2.** Results of survey of families/caregivers and educators/staff.

| **Prior to/without baseline screening** |  | **After baseline screening** |  |
| --- | --- | --- | --- |
| Proportion self-reporting (%) |  | Proportion self-reporting (%) |  |
| ***Educators/staff*** |  |  |  |
| Not comfortable | 50 |  |  |
| Somewhat comfortable | 30 | Not reassured | 5 |
| Neutral | 7 | Neutral | 13 |
| Mostly comfortable | 10 | Reassured | 53 |
| Very comfortable | 2 | Very reassured | 29 |
| ***Families/caregivers*** |  |  |  |
| Not comfortable | 18 |  |  |
| Somewhat comfortable | 31 | Not reassured | 1 |
| Neutral | 12 | Neutral | 12 |
| Mostly comfortable | 27 | Reassured | 41 |
| Very comfortable | 12 | Very reassured | 46 |

Perceived benefits of the screening program for families/caregivers included outbreak prevention (91%), increased opportunity for in-person learning (88%), increased safety for educators/staff and their families (80%) and for students and their families (80%), and a reminder to follow safety protocols (46%). Perceived detriments of the screening program for families/caregivers included cost (35%), false sense of security (30%), stigma (6%), privacy (5%), burden to collect samples (4%), impact on family if student tests positive (4%), and impact of teachers needing to quarantine (4%); 47% reported no perceived detriments.

Assay costs averaged $10/individual screened/week, including both pooled screening and follow-up individual diagnostic testing. Community fundraising supported the monetary costs of the pilot screening program. Staff and volunteers from across the district participated in implementation and operation of the screening program. Community education and outreach via webinars, website information, and weekly messaging for all stakeholders (educators, staff, families, caregivers, students, and local public health agencies) was crucial for the success of the program. The district technology department developed and maintained a Google™ site for registering barcodes for each student and sample, requiring 4 hours for initial development and 1 hour/week for maintenance. Approximately 10 hours per week of parent volunteer time was needed to order and assemble saliva collection kits, apply and track barcodes, collect and transport samples, and maintain data for billing. and review and address results. Bus and van drivers, not driving on Wednesdays due to fully remote learning, spent a total of 54 hours/week assembling test kits; on testing days, they also collected samples at individual schools and transported them to a central location for processing and shipping.

School nurses reinforced correct collection technique, sorted and prepared samples for shipping, and maintained records of participation. Nurses spent an average of 4 hours per confirmed SARS-CoV-2 case (range, 1-6 hours) to conduct contact tracing, provide education/support, and collaborate with the health department. In addition, the Director of Nursing spent at least 20 hours per week on result follow-up, management of positive cases, data entry, and program administration. Nursing, administration, and parent volunteer staff also dedicated additional evening and weekend hours consulting with other public-school districts implementing screening programs.

## DISCUSSION

During the COVID-19 pandemic, preK-12 schools can offer safe in-person learning with infrequent in-school transmission, as long as mitigation measures are implemented uniformly.^16,17^ Weekly screening of educators/staff and middle and high school students can successfully detect asymptomatic cases and in-school transmission events. Over 18 weeks, this screening program detected 37 of the 126 total positive tests among students and staff engaged in in-person learning, and in-school transmission was rare. These findings are consistent with data from Australia, the UK, Norway, Germany, and Italy,^7–9,18,19^ as well as emerging data from K-12 schools across the US.^10,16,17,20–22^ However, rare in-school transmissions did occur associated with breaks in mitigation protocols, consistent with data from Israel, Utah, and Mississippi.^23–25^ The information gained from the screening program was valuable not only in understanding current prevalence of SARS-CoV-2 in the school community, but also in mitigating risk, by identifying a small number of in-school transmissions before additional in-school transmission occurred, and allowing investigation and improvement in potentially associated practices.

Although the risk for severe COVID-19 is relatively low in children, unrecognized or asymptomatic infection in students places their household members at risk for infection.^26,27^ At the same time, there are substantial non-COVID-19-related mental and physical health risks associated with lack of in-person learning.^3,28–31^ Inequities that existed before the pandemic in education and health are exacerbated by school closures, particularly in Black, Latinx, and Indigenous communities and among children experiencing housing and food insecurity.^32,33^ Educator health must also be a key priority; 25-50% of educators have medical conditions or older age that place them at risk for COVID-19 complications, or live with household members with a higher-risk condition.^34,35^ Moreover, teachers and other school staff may have job-related risks that differ from other workplace settings, such as the need for close student interactions over prolonged durations, and shared office space. Developing strategies to maintain workplace safety for educators is critical for schools to operate during this pandemic. These results highlight unique issues for educator safety that extend beyond the usually discussed in-classroom mitigation strategies, including the need for well-ventilated, distanced places for eating and drinking, as well as workspace with adequate distancing between staff. These findings are consistent with SARS-CoV-2 outbreaks observed in other non-school low-risk settings, such as clusters of infection in hospital employees linked to eating and drinking in shared breakrooms.^36,37^

Screening for SARS-CoV-2 in the preK-12 setting, by itself, cannot replace the other layers of protection afforded by the mitigation strategies that have been shown to be effective, including masking, physical distancing, hygiene, and ventilation. Screening can, however, provide an added layer of protection to increase the safety of in-person schooling during the pandemic. Screening programs can detect infections and prevent onward transmission, possibly avoiding larger clusters. Modeling suggests that weekly screening may permit 5-day in-person education at the same total COVID-19 risk as 2-day hybrid education without screening; this effect is larger in the setting of rising community rates.^38,39^

In addition, screening programs can provide data to inform responses to detected cases, e.g., allowing targeted closing of specific classrooms, office spaces, or school buildings while the remaining facilities continue in-person learning/work. In this district, based on global, state, and local experience combined with the data from the screening program, the district dashboard and closure metrics were adapted after the first semester to include these more relevant building-specific metrics, moving away from state and county infection rates as triggers to consider district closure.^40^ Crucially, screening can also provide assurance to faculty, staff, parents and students that mitigation measures are working to prevent spread of infection within schools.

The resources required to implement weekly screening in public schools are substantial. This pilot program cost more than $320,000 in assay costs alone over a period of 18 weeks and required at least 80 hours per week of staff and volunteer time to implement. Many private K-12 schools and universities have been able to implement screening programs; public schools will need extensive state and federal support for screening. While this project was a pilot designed to evaluate the feasibility of screening, it was implemented successfully only because the town had extensive financial resources and availability of private funds. Local, state, and national leaders must recognize that use of private funds is neither a sustainable nor an equitable approach. The cost to implement weekly screening for all K-12 public school students in the US has been estimated at $42.5 billion.^41^ The rapidly improving quality and availability of SARS CoV-2 antigen tests may reduce costs in the future, but accuracy (both sensitivity and specificity) of many of these tests remains lower than PCR.^42^ Use of antigen testing for screening would require extensive governmental oversight and support, including the waiving of the FDA EUA restriction around use for symptomatic people only, a plan for the follow-up of potentially false negative and false positive results with readily available confirmatory testing, and liberalizing rules around collection by non-licensed individuals.^43^

In addition to costs, the logistical challenges of screening in K-12 public schools must be addressed. Important barriers include complex and time-consuming specimen labeling, collection, and transport; notification of faculty/staff and parents of results; and transmission of results to health officials for contact tracing and additional testing. The feasibility of this program was enhanced by the use of saliva samples which were easy to collect at home and the lack of requirement to provide multiple HHS-required data elements for diagnostic testing. The in-laboratory transition from screening testing to diagnostic testing after identification of a positive pair of samples, without the need for an additional specimen, minimized unnecessary isolation of uninfected students and staff.

Participation in screening in most public-school settings is voluntary. Education and outreach to maximize participation will permit the greatest benefit and return on investment for screening programs. Resources describing barriers and facilitators to participation should be publicly shared by districts with successful screening programs.^44,45^ Although these challenges are substantial, they are not impossible to overcome, and are critically needed to protect students, educators, school staff, and the wider communities in which schools operate. Vaccination is anticipated for educators/staff by mid-2021, but students, especially those younger than 12, are likely to remain unvaccinated for some time. The role for screening programs in protecting student and educator/staff health, as well as in providing data and reassurance, is anticipated to persist.

As anticipated for many public-school districts implementing screening, our results are limited to a single town. In addition, because screening testing was not mandatory, the results from the participating population might not represent the entire school community.

In summary, schools provide an essential function in society, yet they present distinct challenges to operate safely during the COVID-19 pandemic. Screening programs can detect the presence of asymptomatic SARS-CoV-2 in schools and, when applied alongside other proven mitigation strategies, allow public schools to continue to operate safely despite rising community rates. Local, state, and federal coordination and support for screening programs will be essential to ensure that all children are provided an equal opportunity for public education and that educators are ensured of workplace safety.

## Data Availability

All data are available on request.

## ACKNOWLEDGMENTS

We gratefully acknowledge the students, families, faculty and staff of Wellesley Public Schools; the Wellesley Education Foundation; the Safer Teachers, Safer Students Testing Collaborative; the Wellesley Board of Health; and the nursing staff and bus/van drivers of the Wellesley Public Schools. We also thank Alyssa Amick, MPH, for assistance in manuscript preparation.

